# Descriptive and prognostic value of a computational model of metastasis in high-risk neuroblastoma

**DOI:** 10.1101/2020.03.26.20042192

**Authors:** Sébastien Benzekry, Coline Sentis, Carole Coze, Laëtitia Tessonnier, Nicolas André

## Abstract

High Risk Neuroblastoma (HRNB) is the second most frequent solid tumor in children. Prognosis remains poor despite multimodal therapies. Mathematical models have been developed to describe metastasis, but their prognosis value has yet to be determined and none exists in neuroblastoma.

We established such a model for HRNB relying on two coefficients: *α*(growth) and *μ* (dissemination). The model was calibrated using diagnosis values of primary tumor size, lactate dehydrogenase circulating levels (LDH) and the meta-iodo-benzyl-guanidine (mIBG) SIOPEN score from nuclear imaging, using data from 49 metastatic patients treated according to the European HR_NBL1 protocol.

The model was able to accurately describe the data for both total tumor mass (LDH, *R*^2^ > 0.99) and number of visible metastasis (SIOPEN, *R*^2^ = 0.96). Statistical analysis revealed significant association of LDH with overall survival (OS, p=0.0268). However, clinical variables alone were not able to generate a Cox-based model with sufficient prognosis ability (p=0.507). The parameter *μ* was found to be independent of the clinical variables and positively significantly associated with OS (p = 0.0175 in multivariate analysis). Critically, addition of this novel computational biomarker to the clinical data drastically improved the performances of predictive algorithms, with a concordance index in cross-validation going from 0.755 to 0.827. The resulting signature had significant prognosis ability of OS (p=0.0353).

Mechanistic modeling was able to describe pathophysiological data of metastatic HRNB and outperformed the predictive value of clinical variables. The physiological substrate underlying these results has yet to be explored, and results should be confirmed in a larger cohort.

**Significance:** A mechanistic mathematical model of metastasis in high risk neuroblastoma is able to describe clinical data and provides a numerical biomarker with superior predictive power of overall survival than clinical data alone.

## Introduction

Neuroblastoma is the second solid tumor in children (8-10% children cancers in USA and Europe) with a median age at diagnosis around 2 years (1,2). Neuroblastoma is responsible of almost 15% of childhood deaths by cancer (3). Neuroblastoma is a quite heterogenous disease at clinical, histological and biological levels (4). Consequently, its prognostic spectrum is also wide (5). The International Neuroblastoma Risk Group (INRG) proposed in 2009 a classification model depending of cancer data (dissemination of neuroblastoma, histology, grade of tumor differentiation, genetic abnormalities such as MYCN amplification (6) and age (3,7)). Therefore, neuroblastoma is divided into 3 risk-groups : Low, Intermediate and High Risk Neuroblastoma (HRNB), which display different survival rates. For patients treated according to the International Society for Paediatric Oncology European (SIOPEN) recommendations, 5-year overall survival is more than 90% for the first group thanks to minimal therapeutics (surgery and/or chemotherapy or simple overseeing), 60 to 80% for the second (5) and < 50% for the lasts group, representing nearly 50% of patients (3,8–11), despite intensive multimodal treatments. Furthermore, patients progressing during or after initial response to induction have a dismal 5-year event-free survival (<20% for patient with early progressive disease (12,13)). For these refractory patients, current therapeutics are unsatisfactory and new treatments or therapeutic strategies are needed.

As early as 1964 (14), efforts have been made to develop mathematical models to assist cancer research (15). Their aim was to understand multiple biological processes involved in cancer and to propose rational tools for the design of therapeutic drug regimen (16,17). Three main types of mathematical models can be distinguished. On one hand, highly complex, multiscale models try to integrate as much of the biology as possible, ranging from intra-cellular molecular processes to systemic interactions at the whole organism level (18). This approach requires many parameters and consequently the models are often impossible to reliably calibrate for clinical purpose. On the other hand, purely statistical models and artificial intelligence techniques rely on agnostic algorithms that try to learn patterns directly from the data (19,20), with applications mostly in genomics (21) and radiology (22), but rarely in clinical oncology. In between, mechanistic or semi-mechanistic models seek to describe only the main determinants of a cancer disease, for a given purpose (e.g., prediction of survival from tumor kinetics (23), understanding (24,25) or prediction (26,27) of metastatic relapse).

To our knowledge no mechanistic model has yet been established and validated for clinical neuroblastoma. In this study, we define such a model for high-risk neuroblastoma to describe the metastatic burden at diagnosis, using two coefficients: a patient specific parameter *μ* for the dissemination process and a patient nonspecific parameter *α* for the growth process. The model was built and calibrated using clinical, biological and radiological data from a monocentric cohort of 49 patients with HRNB treated according to the HRNBL1 protocol (9). We then evaluated the prognostic value of the individual parameter *μ* and tried to identify Ultra High-Risk patients.

## Material and methods

### Ethics statement

Authorization to perform the study was obtained at APHM (Public Assistance of Marseille’s Hospitals) Health Data Access Portal (number request 32PTJ5)). We respect the Informatic and Liberty Law (1978) for the use of data. All parents and patients when appropriate gave consent to participate in the study.

### Data collection

Our population is made of 49 patients with HRNB (see stratification algorithm in Table S1) treated according to the HRNBL1 protocol recommendations (9), in the paediatric hematology and oncology unit of the children hospital of the University Hospital of Marseille (AP-HM) between 11/26/2007 and 08/30/2018. Entry date was the date of diagnosis. For survival analyses, end date was either the date of patients’ death or the date of last news. Inclusion criteria were the inclusion criteria of the HRNBL1 protocol (9) (see Figure S1). Briefly, induction chemotherapy with “rapid COJEC” or “modified N7 induction” is given for 10 weeks, followed by surgery when considered possible, then myeloablative chemotherapy with hematopoietic peripheral stem cell transplantation. Treatment is then completed with radiotherapy and maintenance therapy with immunotherapy (anti-GD2 ± IL2 therapy) and retinoic acid for 6 months.

Data were gathered from Personalized Computerized Folders (PCF) by the Axigate platform used in AP-HM, which includes neuroblastoma risk factors such as age at diagnosis, lactate dehydrogenase (LDH) – correlated to the total tumor volume (6,9,28,29) – or MYCN amplification, researched by polymerase chain reaction from peripheric blood and/or from primary or metastatic tumor tissue at diagnosis. The dataset is available in the supplementary file S1.

### SIOPEN scoring (Figure S2)

The meta-iodo-benzyl-guanidine (mIBG) is known to bind to neuroblastoma cells using iodine 123 (I123) (30) and mIBG scintigraphy is consequently used to evaluate the extent of the disease, in agreement with INRG recommendations. Indeed, mIBG binds to almost 90% of neuroblastomas (31) in primary tumors, but also bone, bone marrow (32) or even soft tissues with a high sensibility (85-94%) (33). We used a semi quantitative SIOPEN score that was elaborated to predict extension and severity of the disease (34). A high score has been shown as pejorative but no reproducible cut off has not yet been found (31,34). We established the SIOPEN score with the PCF data using the Centricity imaging software, or by retrospective double scoring scintigraphy with an experiment nuclear physician (*LT*).

### Tumor characteristics

Location and size of primary tumors was evaluated using radiological reports performed at diagnosis. Forty-five patients (91.8%) had scanner, 11 patients (22.4%) magnetic resonance imaging (MRI). Primary tumor volumes were estimated by the formula: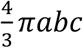 with *α* half the largest axis, *b* half the medium axis and *c* half the smallest axis of an ellipsoid tumor.

Location and number of visible metastases were retrieved from mIBG scintigraphy imaging. Metastases locations were recorded in imaging interpretation. In addition, bone marrow metastases were searched by performing myelograms and bone marrow biopsies.

Date of best treatment response was recorded according to the International Neuroblastoma Response Criteria (INRC) (35). Date of relapse was recorded as the date on which unfavorable evolution of the disease was highlighted by radiology (scanner and/or MRI) and nuclear imaging (positron emission tomography and/or mIBG I123 scintigraphy). We defined Ultra-High-Risk (UHRNB) patients as relapsing or progressing within 18 months after diagnosis.

## Mathematical model

### Definition

The mathematical model was based on a previously published framework (25,26,36), which allows to simulate a cancer disease at the organism scale, including growth of the primary tumor (PT) and birth and growth of secondary lesions (Figure 1). We assumed growth of both the primary and secondary tumors to follow an exponential law:

**Figure 1:**
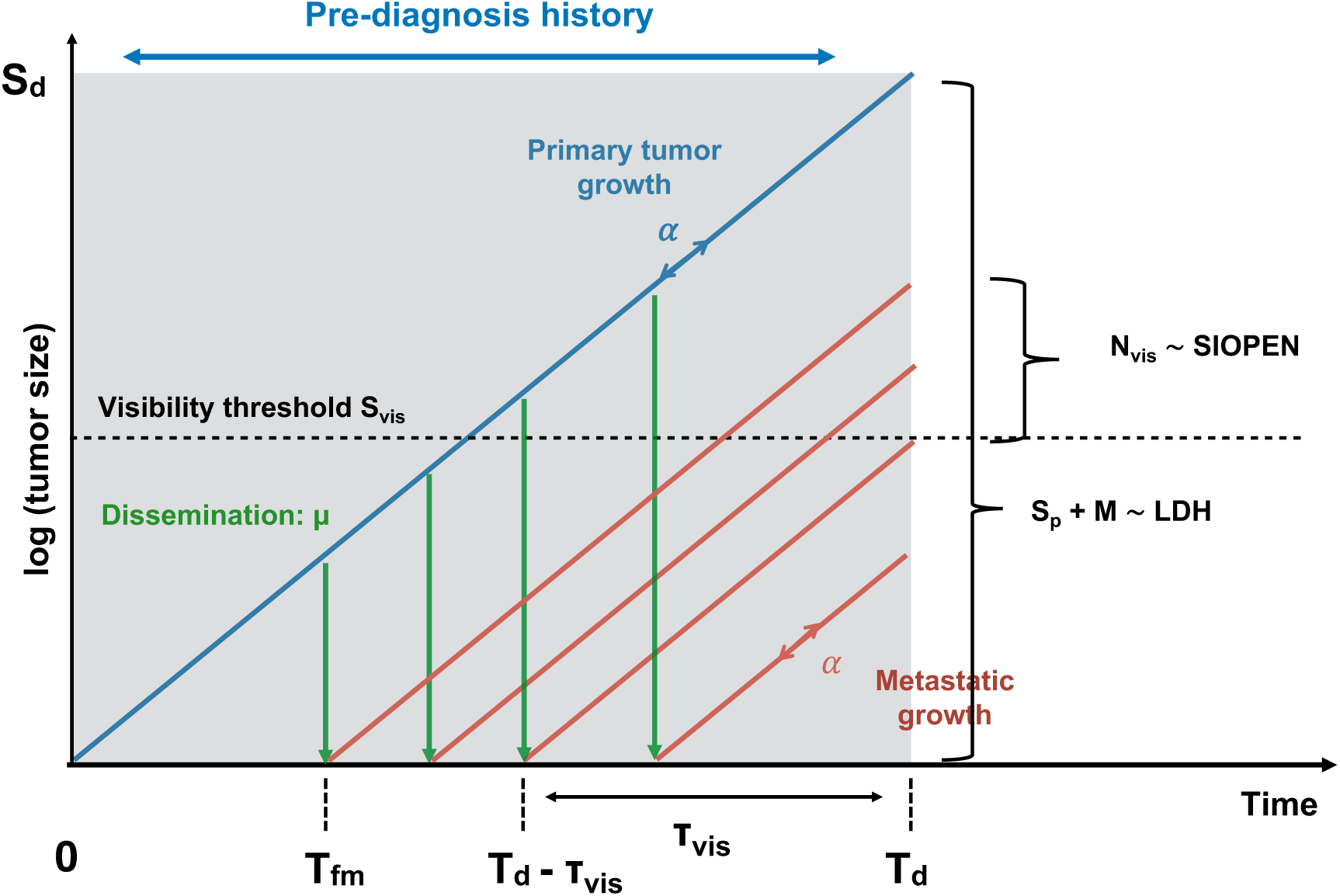
Schematic of the mathematical model. Primary and secondary tumors are assumed to have exponential growth kinetics governed by a proliferation rate *α*. Dissemination of metastasis is controlled by the parameter *μ*. From these and primary tumor size at diagnosis *S*_*d*_, the primary tumor age *T*_*d*_ can be computed and simulations of the natural history can be performed. Adjunction of a visibility threshold *S*_*vis*_ results in predictions of the number of visible metastases *N*_*vis*_ and total cancer mass (primary + secondary tumors) *S*_*p*_ + *M*. These are respectively compared to the SIOPEN score and lactate dehydrogenase level. The time of birth of the first metastasis is denoted *T*_*fm*_ and the time to reach *S*_*vis*_ from one cell τ_*vis*_. Note that the number of visible metastases at time *T*_*d*_ is the total number of metastases at time *T*_*d*_ − *τ*_*vis*_.

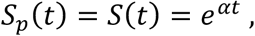

where *S*_*p*_(*t*) and *S*(*t*) denote the sizes of a primary and a secondary tumor (expressed in number of cells), starting from one cell at time *t* = 0. The parameter *α* denotes the proliferation rate. Assuming a metastasis birth rate proportional to the PT size with parameter *μ*, the number of metastasis at time *t* is given by (25):

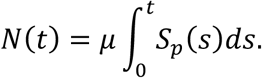

The parameter *μ* corresponds to the per day probability for each cell of the PT to spread and establish a distant metastasis. The total metastatic burden (total number of metastatic cells in the organism) is given by (26):

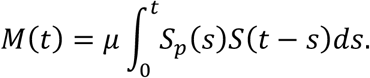

Visible metastases at time *t* (i.e. metastases with size larger than a visibility threshold *S*_*vis*_) are the ones that were born early enough to have reached *S*_*v*i*s*_ at *t*, that is, before *t* − τ_*vis*_, where τ_*v*i*s*_ is the time to reach *S*_*v*i*s*_ (see Figure 1). This time is given by 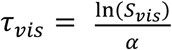 and the number of only visible metastases can then be computed as:

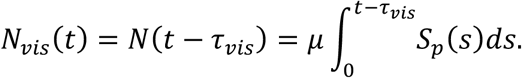

The visibility threshold *S*_*v s*_ is considered a patient-specific model parameter. Numerical simulations of M were performed using the fast Fourier transform algorithm as implemented in the *scipy* python package (python 3.7), exploiting the convolution structure of the equation (37).

For forward simulations of the model, a discrete version was employed with initiation time *T*_*i*_ and size *S*_*i*_ of the i-th metastasis given by:

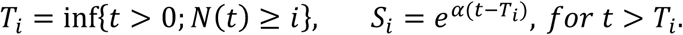

### Calibration

Cell doubling times (CDTs) in neuroblastoma was a prerequisite to estimate *α*. We searched PubMed for studies related to CDT, as well as CDTs from a database of commercial cell population (supplementary file S2). All cell lines were obtained from human patients and CDTs were established in vitro. Of the 73 strains studied, 15 were excluded due to a lack of knowledge regarding possible exposure to chemotherapy.

Median CDT was 48h (20 - 258h). We then fixed 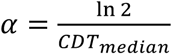. This further allowed us to compute an estimation of the PT age (or time of diagnosis, *T*_*d*_):

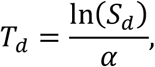

where *S*_*d*_ is the size of the PT at diagnosis. This last quantity was derived from three diameters obtained from computed tomography imaging, which allowed computation of the PT volume assuming ellipsoidal shape. This volume was converted into a number of cells using the standard assumption of 1 mm^3^ ≃ 10^6^ cells (38).

For each patient, two quantitative measurements were used to compare the metastatic model to the data: the SIOPEN score and the LDH blood level. The former was assumed to be a surrogate of the visible number of metastasis and the latter to represent the total cancer burden in the organism (PT + metastases, Figure 1). Denoting with *i* superscript the quantities that depend on individual *i*, we thus assumed:

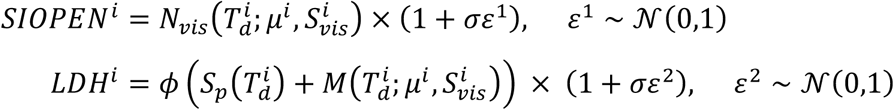

which expresses a proportional error model for the observations with standard deviation *σ* = 0.1, corresponding to a 10% measurement error. The parameter *ϕ* represents a patient non-specific conversion coefficient between number of cells and LDH (expressed in units per liter, UI/L). Maximization of the log-likelihood for the expression above leads to minimization of the following objective function:

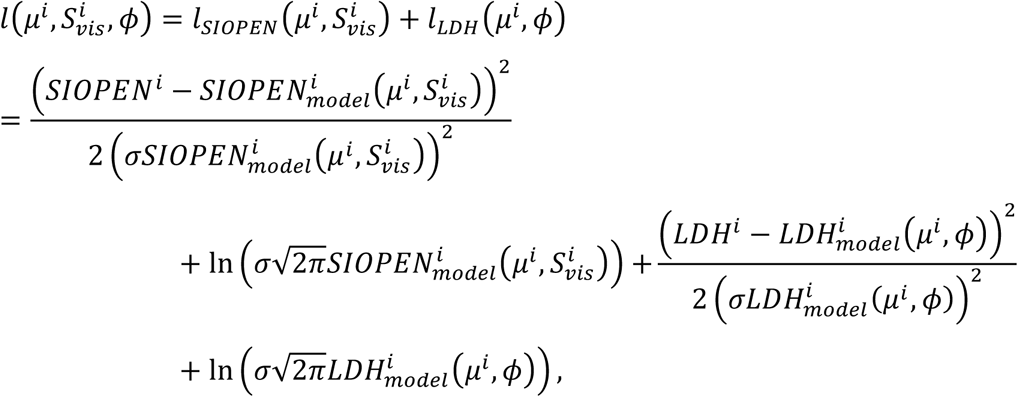

With

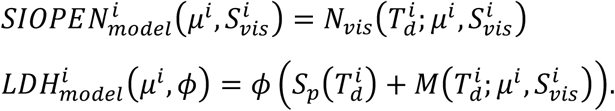

The parameter *ϕ* was arbitrarily fixed to 10^−9^ UI/L/cell, from preliminary simulations. Minimization was implemented using the Nelder-Mead algorithm of the *minimize* function of the *scipy* python package (python 3.7).

### Statistical and predictive analysis

Due to ranges spanning several orders of magnitude, individual values of LDH levels and the mathematical parameter *μ* were log-transformed beforehand. Association between clinical variables and/or the individual mathematical parameter ln *μ* with progression-free survival or overall survival was assessed using log-rank tests for dichotomized groups, as well as univariate and multivariate proportional hazard Cox regression models for continuous covariates. The *lifelines* python package was used to fit the models. Resulting models were evaluated for their predictive power by computing the mean of Harrell’s c-index (39) during a ten-folds cross-validation procedure. It is the equivalent of the area under the ROC curve for survival analysis (proportion of correctly predicted pairs of subjects). Specifically, we first selected the variables below a p=0.2 significance threshold in univariate analysis. Then multivariate Cox models including only these variables were trained on the cross-validation learning sets, and the c-index was computed on the corresponding test sets. For construction of prognosis scores, Cox models were trained using the selected variables on the entire data set. For a patient *i* with covariates *x*, the score was defined by *β*^T^*x* with *β* the vector of the Cox coefficients. Calibration of these Cox models was assessed using calibration plots at the median survival value. These bin patients according to predicted survival and compare survival predictions with the Kaplan-Meier estimates of the patients within the bin (*calibrate* function of the *rms R* package).

## Results

### Description of the cohort

#### Patients and tumor characteristics (Table 1)

Forty-nine patients were included in our cohort. Two girls of 26 and 11 months diagnosed with low risk neuroblastoma were included due to early progression. The MYCN status for both patients was negative at diagnosis but changed for the younger one at relapse. We excluded 4 patients for the construction of the mathematical model as the date of inclusion in the HRNBL1 protocol was delayed when compared with the initial diagnosis (pre-treatment with other off-protocol chemotherapy types and one 36 months-old girl with a metastatic, MYCN negative esthesio-neuroblastoma whose size could not be estimated).

**Table 1:**
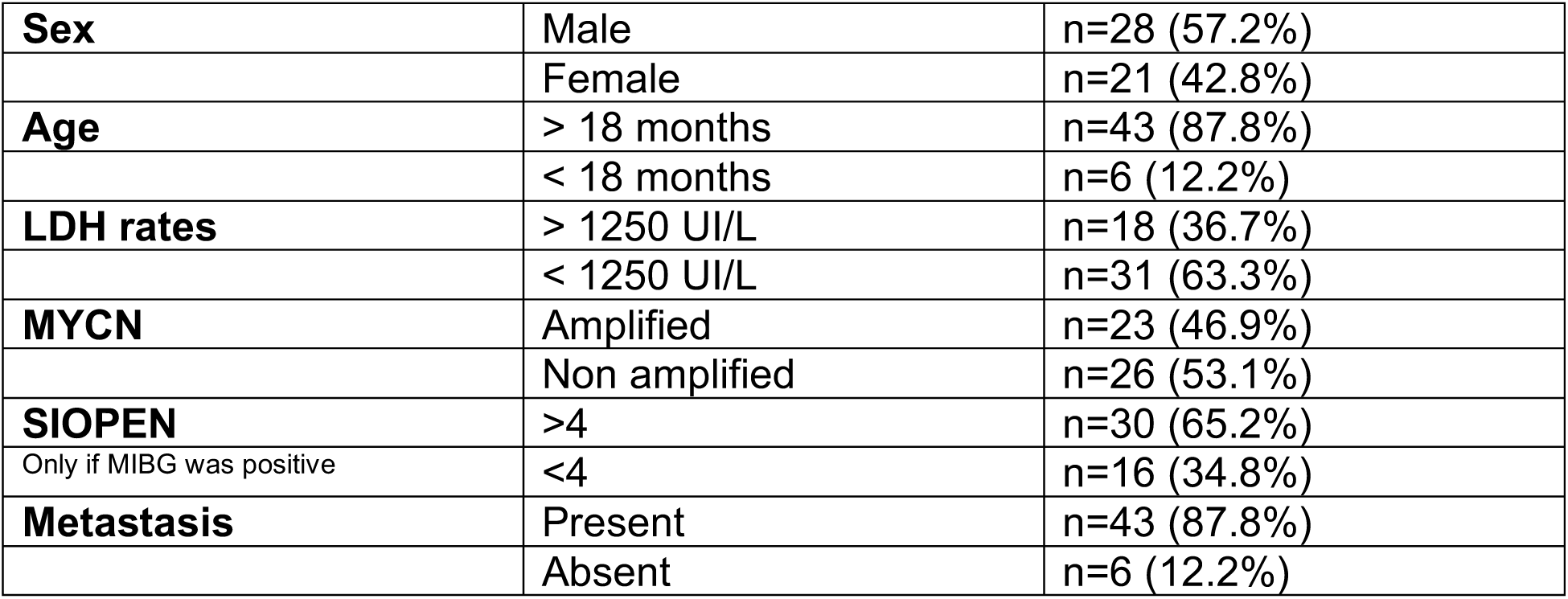
Patients characteristics

Median age was 36 months (range 11-140). LDH levels at diagnosis were high with a median of 842 UI/L (302-22022), compared with laboratory normal values < 300 UI/L. Metastases were present for most patients (91.8%) and SIOPEN scores were overall high (median 27 (0-60)). Three patients who had a negative mIBG (no fixing primary tumor on scintigraphy) but PET-visible metastases were excluded. All patients underwent bone marrow aspirates and/or biopsies.

Location of primary tumors was adrenal for 55,1% patients (n=27) and abdominal for 34,7% (n=17). Details are given in Figure S3A. Primary tumor median volume was 272 cm^3^ (range 0.5 – 2 266 cm^3^). Locations of metastases are detailed in Figure S3B. The most frequent metastatic site was bone marrow (n=38, 77.6%).

### Patients outcome

All patients were evaluable for response to induction chemotherapy. Twenty-three patients had complete response (46.9%), 24 partial response (49%) and only 2 had stable disease (4.1%). However only 20 patients did not ultimately progress (40.8%). Among those who progressed (59.2%), 25 died (51% of whole initial cohort). Details of patients survival are showed in Figure 2. The median survival without progression was 29 months. At 3 and 5 years, progression-free survival rates were 44.1%, and 29.1%, respectively. Median overall survival (OS) was 43 months. At 3 and 5 years, OS rates were 55.8% and 38.9%, respectively.

**Figure 2:**
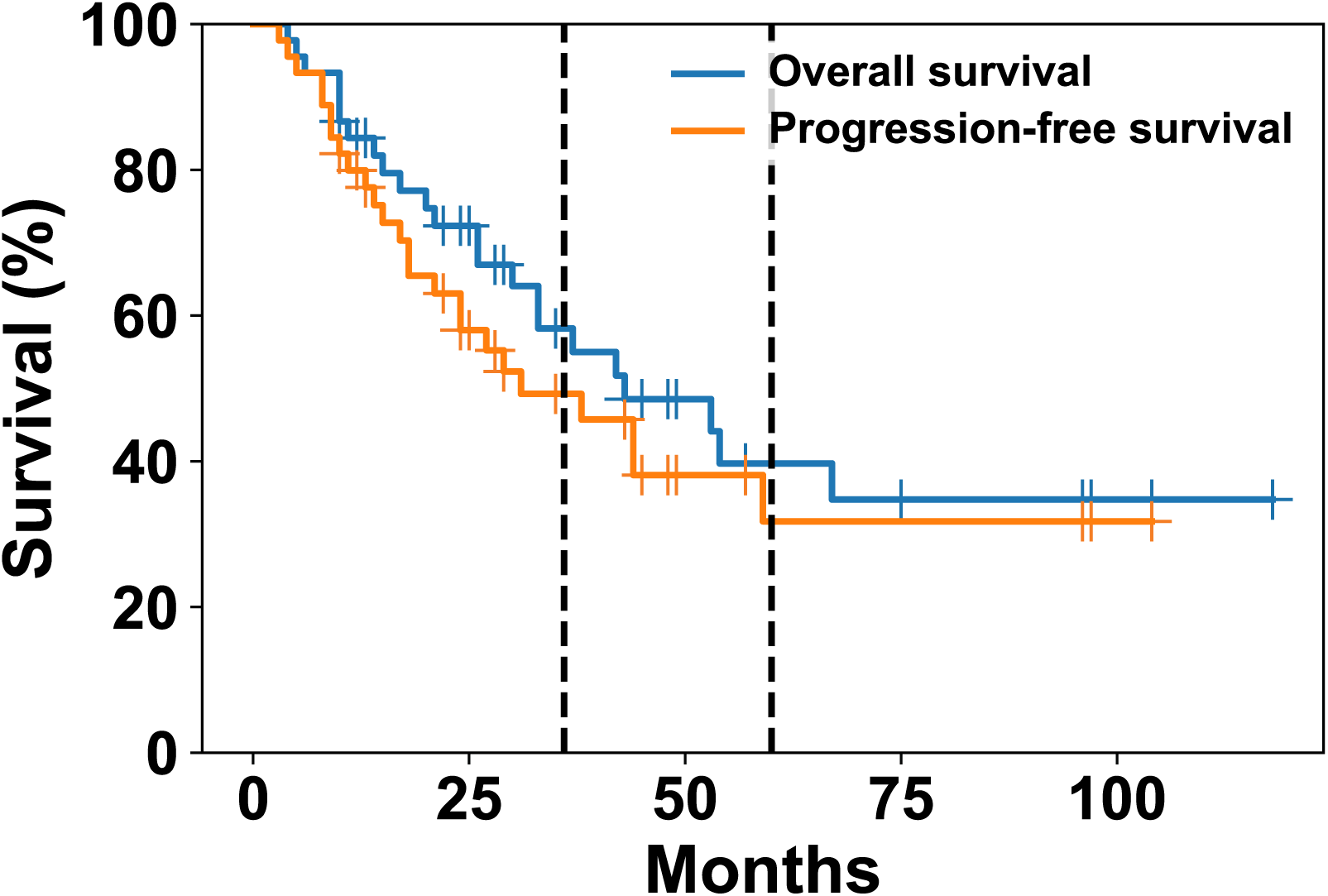
Overall and progression-free survival.

### Descriptive power of the mathematical model

To describe the metastatic burden of HRNB patients, we developed a semi-mechanistic modeling approach whereby the metastatic process is reduced to two main phenomena: growth and dissemination (Figure 1). Growth was assumed to be exponential and the dissemination rate to be proportional to the primary tumor size, with a proportionality factor *μ*. To rely to the data and estimate *μ*^*i*^ in a given patient *i*, we assumed that the LDH level was a surrogate of the total tumor mass, whereas the SIOPEN score reflected the number of visible metastases (Figure 1). We also used the primary tumor size at diagnosis to infer the age of the tumor and simulate the pre-diagnosis history of the disease. The model was able to accurately reproduce the SIOPEN score and LDH levels (Figure 3A-B, *R*^2^ = 0.96 and > 0.99, respectively). Interestingly, The parameter ln *μ* revealed no correlation with either the log(LDH) (R = 0.25) or the SIOPEN (R = 0.201, Figure 3C), suggesting independent added value of this parameter – possibly informative of progression or survival – as compared to the data alone.

**Figure 3:**
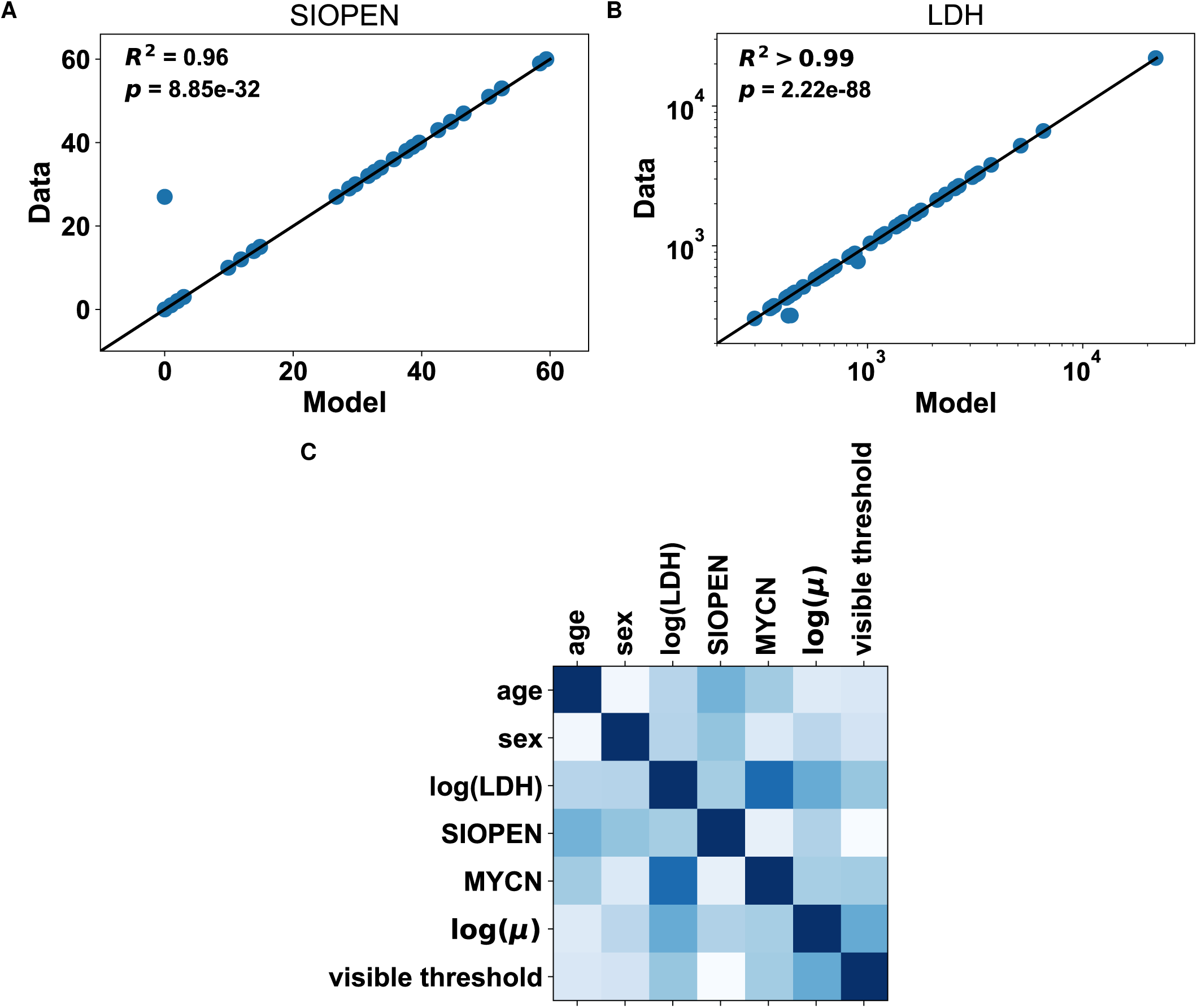
Descriptive power of the mathematical model. A. Fit of the SIOPEN data. Solid line is the identity line. B. Fit of the LDH data. Solid line is the identity line. C.Correlation matrix of all features including clinical variables and (log) of the mathematical parameter *μ*. Level of darkness indicates positive correlation whereas brightness indicates negative correlation.

### Simulations of the natural history of HRNB

Statistical inference of the model parameters *μ*^*i*^ and 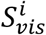 allowed to perform simulations of the predicted natural history of the disease. Representative patients are shown in Figure 4A (see Figure S4 for all patients). These highlight the high inter-individual variability, well captured by the mathematical model (Figure 3). As a general observation however, we predicted that once initiated, metastatic dissemination then experienced a “burst”, with multiple metastasis born in a short period of time (see patients 1, 16, 24, 25 and 30 in Figure 4A). Nevertheless, the time *T*_*fm*_ between the first cancer cell and birth of metastasis was variable, from almost simultaneous to cancer initiation (e.g. patient 24, 0.42 days) to one month (e.g. patient 25, 34.4 days), see Figure 4B. The predicted age of the PT was less variable (median 76 days, range 58.1 – 82.1 days).

**Figure 4:**
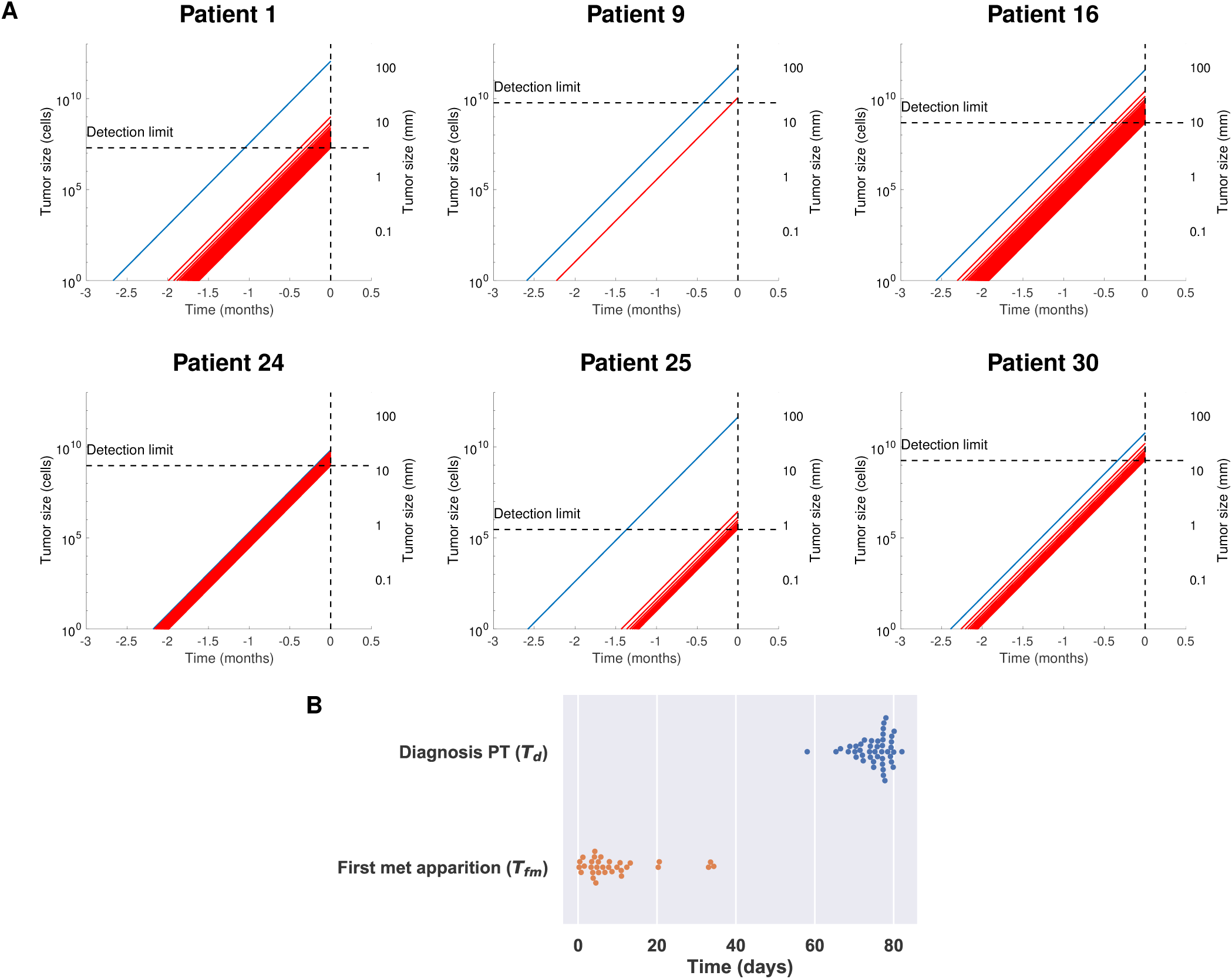
Mechanistic simulations of the pre-diagnosis history of high-risk neuroblas-toma patients. A mechanistic model was calibrated using patient data of SIOPEN score (number of visible metastases) and LDH levels (total cancer burden), which resulted in individual values of the model parameters *μ* and *S*_*vis*_. These values were used to simulate the pre-diagnosis natural history of the disease. A. Simulations of the primary tumor (blue) and metastases (red) growth kinetics, for representative patients. **B**. Distributions of the times of the primary tumor (PT) diagnosis (age of the PT, *T*_*d*_) and birth of the first metastasis (*T*_*fm*_).

## Prognostic analysis

### Analysis of classical prognosis factors

Using log-rank tests for dichotomized groups (Figure S5), no significant difference was found in progression-free survival (PFS) for gender (p = 0.219), MYCN status (p = 0.354) or age (p = 0.825 with, 18 months cut-off). For LDH and SIOPEN, separation at the median value or from literature thresholds (9,34) was not able to significantly discriminate patients and statistical significance was only reached for extreme values, respectively at the 80^th^ percentile (2,375 UI/L) and the 90^th^ percentile (46.2) (Figure S5). In univariate Cox regression analysis, only the LDH level was associated with PFS (Hazard Ratio (HR) 1.6 (95%CI: 1 – 2.56), p=0.05). For analysis of the predictive power, univariate analysis selected the variables LDH and SIOPEN at a p < 0.2 threshold. The mean c-index in cross-validation was 0.603 for PFS.

Similarly, no significant difference was found in overall survival (OS) for gender (p = 0.198), MYCN status (p = 0.181) or age (p=0.527). Again, a significant difference in OS was found only for at the 80^th^ and 90^th^ percentiles of LDH levels and SIOPEN score. Using univariate Cox regression, only the LDH rate was significantly associated with OS (HR 1.74 (95%CI: 1.07 – 2.84), p=0.0268), which was confirmed in multivariate analysis (Figure 5A). The predictive model selected the variables LDH and MYCN status as having p < 0.2, which resulted in a c-index of 0.755. However, these variables were not sufficient to yield a Cox score able to significantly discriminate patients (p=0.507, Figure 5B).

**Figure 5:**
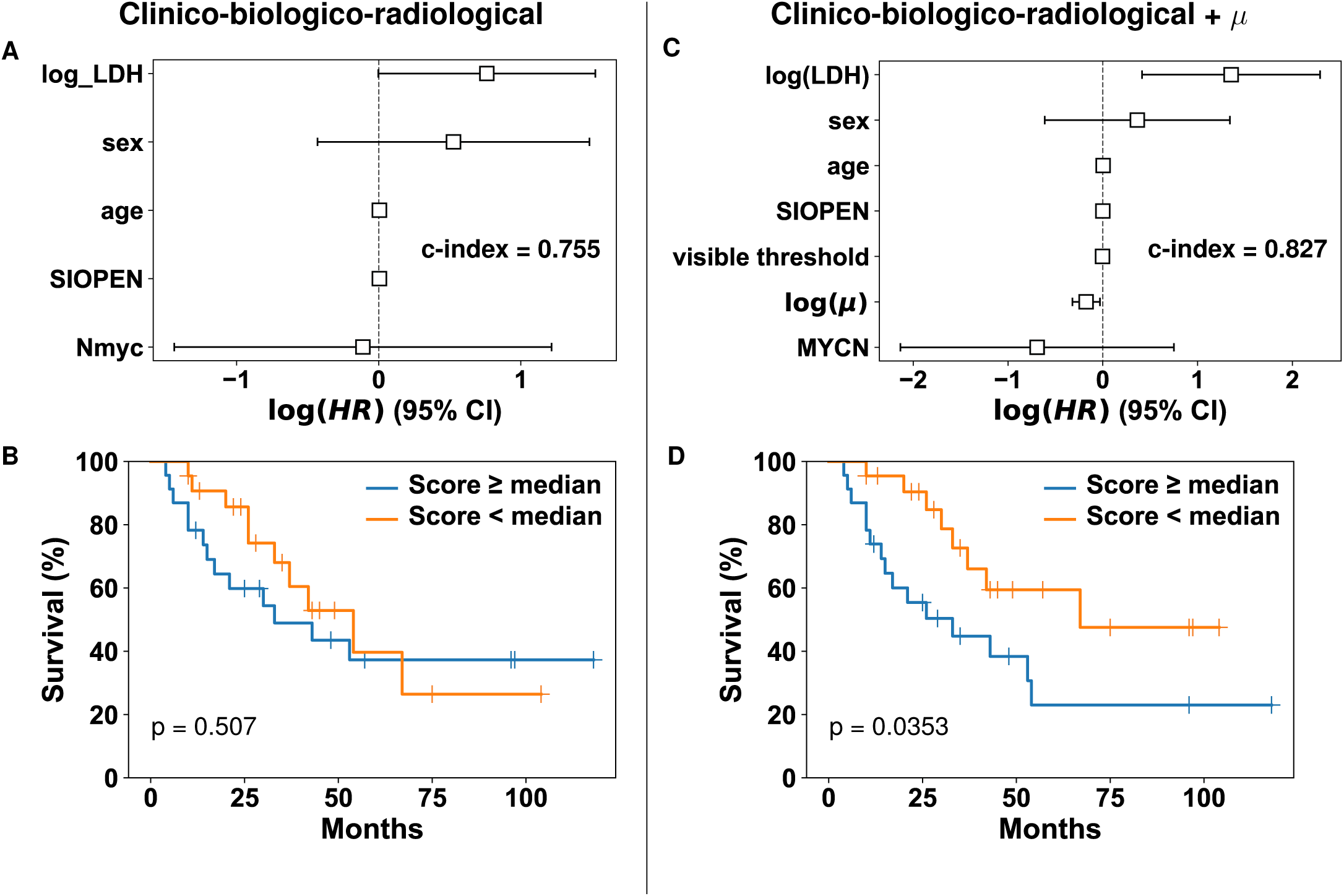
Prognosis of overall survival. Using multivariate Cox regression, we compared the association and prognostic value of clinical variables alone (left) or complemented with the mathematical biomarkers (right). A. Hazard ratios and 95% confidence intervals of the clinical variables in multivariate Cox regression. The reported c-index corresponds to the one obtained in cross-validation using variables selected at p < 0.2 in univariate analysis (log(LDH) and MYCN). B. Separation of patients according to the Cox score predicted from the selected clinical variables (log(LDH) and MYCN). C. Same as A. with log(*μ*) and visible threshold *S*_*vis*_ as additional variables. D. Same as B. with log(*μ*) as additional variable in the model.

### Added value of the mechanistic model

Neither *μ* nor *S*_*v s*_ were significantly associated with PFS (p = 0.475) in log-rank analysis (Figure S5) or Cox analysis. On the other hand, the parameter *μ* seemed to be the quantitative parameter most associated with OS in dichotomized analysis (Figure S6), even if not significant probably due to a crossing of the survival curves. Critically, confirming this observation, *μ* was positively significantly associated with better OS in multivariate Cox regression analysis (HR=0.839 (95% CI: 0.726 – 0.97), p=0.0175, see Table 2 and Figure 5C). Moreover, adding this novel computational biomarker *μ* to the previous significant biomarkers LDH and MYCN strongly increased the predictive power with an updated c-index of 0.827 (+9.54%). In addition, this superior predictive power was further supported by the fact that a novel Cox score, derived from the coefficients of LDH, MYCN and log(*μ*), was now able to significantly separate patients between poor and good prognosis (p=0.0353, Figure 5D). This was also confirmed by better calibration plots when including *μ* vs when not including it (Figure S7).

**Table 2:**
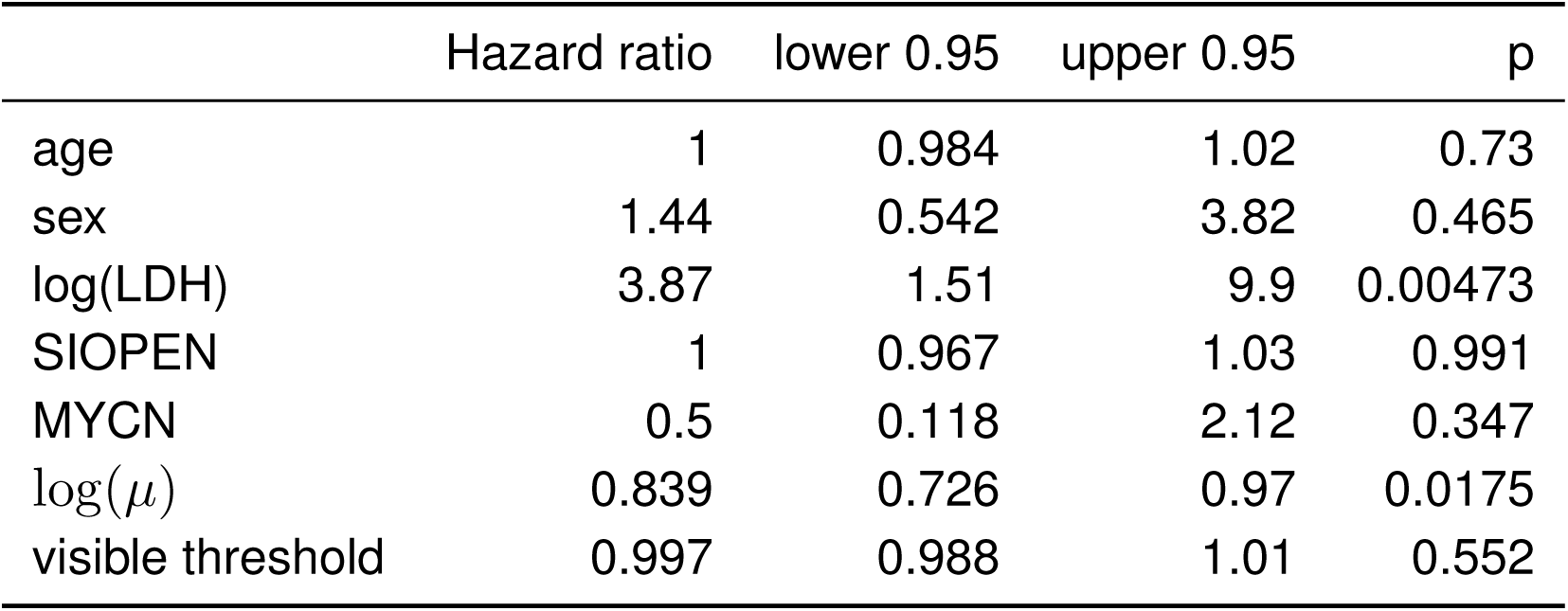
Multivariate Cox analysis of overall survival

## Discussion

We reported here about the development of a mathematical model of metastatic neuroblastoma using a mechanistic approach based on classical risk factors, easy to collect at diagnosis and routinely used by clinicians. Our model could adequately describe total cancer mass represented by LDH levels as well as visible metastases represented by the SIOPEN scores. Beyond this mere descriptive value, our analysis suggested predictive value of a new computational biomarker *μ*, able to significantly better predict outcomes for patients.

Tumor growth is a complex biological process, that includes tumor proliferation, regulation of abnormalities concerning stem cells (40), neoangiogenesis (32,33), microenvironment interactions (40,41), immune interactions with tumor cells and dysregulation immune system (4,41–43). These complex interacting processes are regulated by many genes or epigenetic regulators (44) currently still being investigated. How to model these complex properties remains an open debate. We have used a semi-mechanistic approach relying on both clinical and radiological data to model neuroblastoma growth. Such mechanistic models of metastasis have already been successfully used for different cancer types such as kidney (24), breast (26,27) or lung (25). Although not included here, these models can incorporate the effects of multiple therapies (i.e. surgery, chemotherapy) or tumor-tumor interactions to describe and predict tumor dynamics (26,45,46). Moreover, the limited number of parameters used in these models allows a quick translation to potential clinical applications.

A very limited number of studies have focused on the mathematical modeling of neuroblastoma genesis, growth, or metastatic evolution. Ciccolini et al. (47) have reported a mechanistic model of neuroblastoma growth, using a classical Gompertzian model. Their model was used to optimize gemcitabine metronomic chemotherapy administration but did not intend to model metastatic dissemination. Elsewhere, He et al. (48) coupled a complex vasculature model fitting the dynamic growth of the human neuroblastoma cell line IMR32 in mice and a pharmacokinetics/pharmacodynamics model of bevacizumab, an anti-VEGF agent, to determine the best dosing regimen for this treatment and predict its effectiveness. Kasemeier-Kulesa *and al*. (49) have developed a molecular network model of developmental genes and signaling pathways with a 6 gene inputs logic model, using the discrete Boolean logic, and based on 4 cell states (differentiation, proliferation, angiogenesis, apoptosis). The model was able to predict the stage of the human neuroblastoma SHSY5Y and then the outcome of 77 early stage patients. Recently, Hidalgo et al. (50) modeled the whole cell signaling pathways data linking analysis of different pathways to molecular mechanisms involved in cancer physiopathology and patient survival. They identified numerous pathways implicated in the activation or deactivation of several cell functions responsible of poor outcomes in patients with neuroblastoma by for instance promoting of proliferation and apoptosis inhibition (TP53), angiogenesis (FASLG), or metastasis (THBS1, PTPN11 and cAMP AFDN). All the models proposed above are nevertheless not easily translatable in the clinic.

Alternatively, although not being the mainstay yet, individual molecular profiling has been studied in neuroblastoma (3,4). Several studies explored genome wide associations to predict outcomes for HRNB patients (51,52) but they are not used yet in day-to-day clinical practice. The relevance of the identified markers can also be questioned due to the lack of evidence of a causal relationship (53).

Our mechanistic model is a simplified representation of the metastatic process, reducing it to two basic phenomena: growth (*α*) and dissemination (*μ*). Our findings suggested growth to be exponential and fast, with primary tumors reaching 10 cm tumors in less than 3 months. This contrasts with previous studies where tumor growth kinetics were shown to slow down and deviate from an exponential pattern, following rather a gompertzian pattern (25,54). This is probably explained by the embryonic nature of this cancer, which makes it particularly aggressive. Dissemination was also characterized with a very high value of *μ* (median = 0.0446 metastases/cell/day) in comparison to other adult cancers (e.g. typical *μ* = 2.30 × 10^−l2^ metastases/cell/day in breast cancer). Notably, the inferred values of metastatic sizes at diagnosis and threshold values *S*_*v s*_ fall within biologically realistic ranges (median *S*_*v s*_ = 14 mm), although no a priori metastatic size information was used when calibrating the model. Only numbers of metastases above a parametric threshold (automatically fitted by the model) were used.

The parameter *μ* was found a better prognostic tool than the validated SIOPEN score at diagnosis and can be combined with LDH and MYCN status to predict OS. Interestingly, a high *μ* value is paradoxically an independent and statistically significant factor of better OS in our cohort. This might be explained by two possible hypotheses. First, patients with high *μ* may have an aggressive neuroblastoma with high replicative potential, which may result in a better sensitivity to chemotherapy and therefore better survival. Second, patients with high *μ* have a bigger total cancer burden, but this total mass could result in systemic inhibition of proliferation (46). This would be consistent with the fact that *μ* is a good prognosis factor for OS but not for PFS. Indeed, the visible mass might progress while suppressing the growth of smaller, invisible tumors. Assuming further that death results from the total mass present in the organism and not only the visible lesions could possibly explain why patients who progress are distinct from patients with the largest number of metastases (i.e., largest *μ*). To further confirm or invalidate these hypotheses, further mechanistic insights could be gained by linking *μ* to molecular analyses of the tumor. The micro-environment and more specifically the immune system might also be implicated in slow tumor progression and a host’s tumor long-term control.

The major limitation of our study is the limited (n=45) number of patients included in our analysis, due to the monocentric nature of our cohort. This nevertheless corresponds to all HRNB patients treated at our institution during over a 10 years period. This prevented us to extract a test set from the data before any analysis, on which to evaluate the predictive power of the model a posteriori. Nevertheless, we tested the predictive abilities of *μ* using cross-validation, i.e. on independent sets independent from the learning ones.

In this cross-validation process, the model changes on each fold, thus deciding on the “best” model (i.e. the ones with highest c-index across the 10 folds) still relies on entire data set. Critically, the mean c-index was found higher when using *μ* to predict survival, reaching a very good score (> 0.8). Further research should evaluate the predictive value of this final model on independent data.

## Conclusion

We developed a mechanistic mathematical model, using human data and a limited number of standard risk factors required in the clinic. We have chosen to use prognostic factors that are available at the time of diagnosis, in order to be able to provide upfront alternative therapeutic strategies adapted to the patients most likely to be unresponsive to a conventional high-risk neuroblastoma treatment. The model can reproduce tumor spreading of high-risk neuroblastoma in our patients and also predict patient prognosis, better than clinical variables only. It also led to the creation of a new risk score based on the parameter *μ*, which is associated with better OS outcome in our population. These findings must be confirmed in a larger cohort and the physiological substrate underlying this result should be explored.

## Data Availability

The data will be made available upon acceptance of the manuscript for publication in a peer-reviewed journal

## Supplementary figures and tables

**Table S1:**
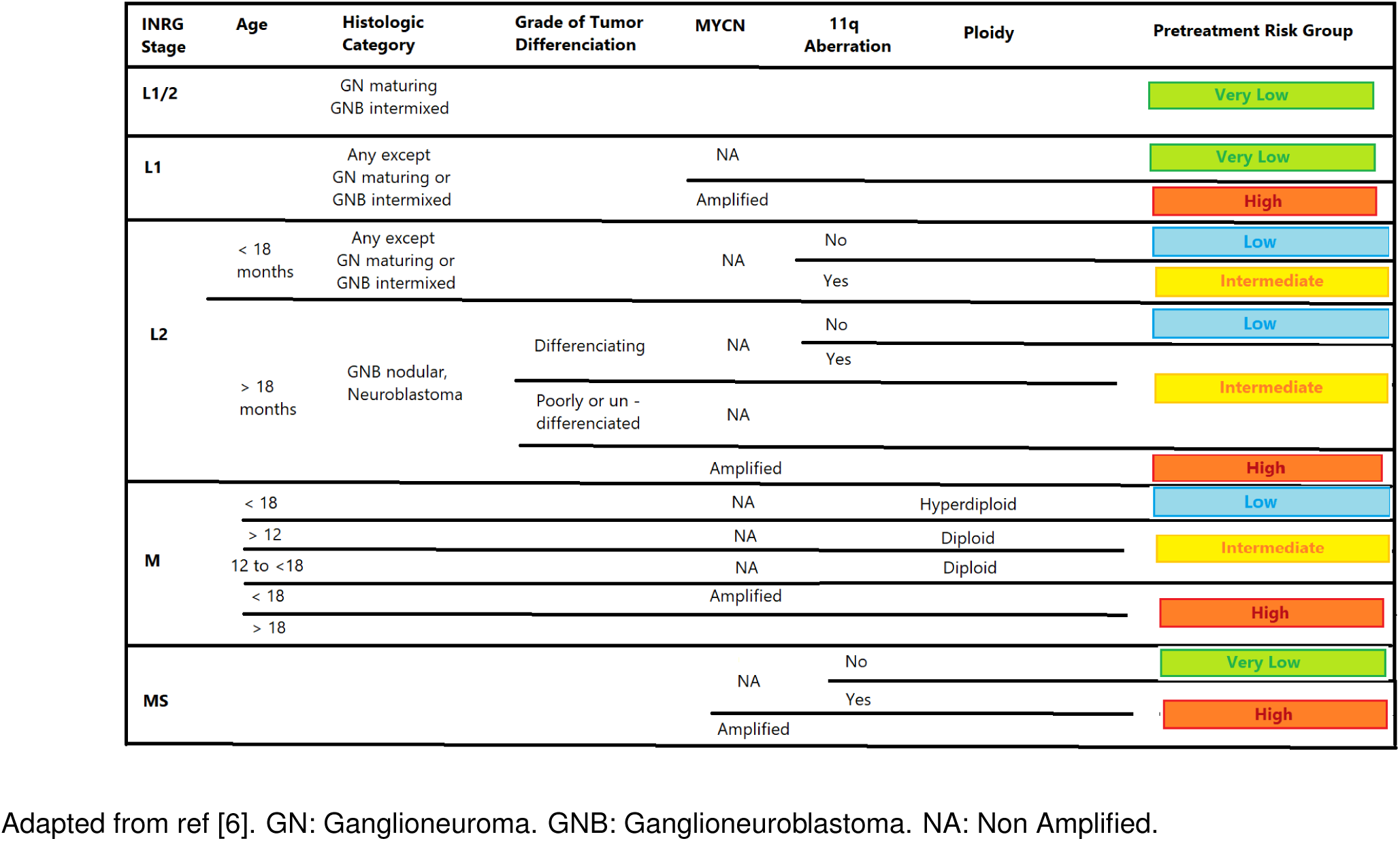
Neuroblastoma classification according to the International Neuroblastoma Risk Group staging system

**Table S2:** Cox analysis of progression-free survival

|  | Hazard ratio | lower 0.95 | upper 0.95 | p |
| --- | --- | --- | --- | --- |
| age | 0.997 | 0.978 | 1.02 | 0.718 |
| sex | 1.15 | 0.461 | 2.85 | 0.768 |
| log(LDH) | 2.42 | 0.985 | 5.92 | 0.0539 |
| SIOPEN | 1.01 | 0.982 | 1.05 | 0.395 |
| MYCN | 0.801 | 0.196 | 3.27 | 0.757 |
| $\log(\mu)$ | 0.902 | 0.79 | 1.03 | 0.13 |
| visible threshold | 0.997 | 0.987 | 1.01 | 0.465 |

**Figure S1:**
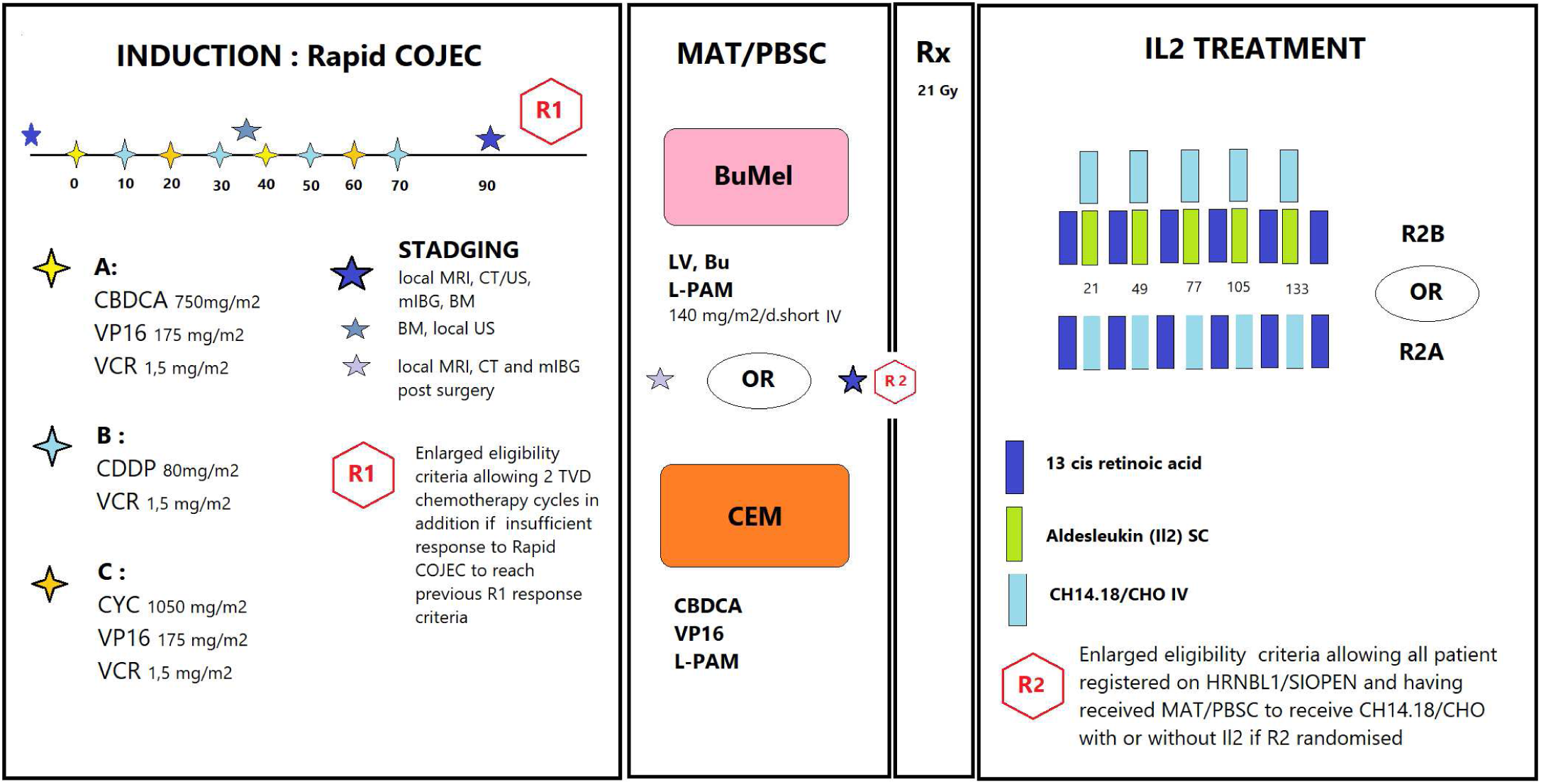
HRNBL1 protocol. Time is in days. Staging: MRI: Magnetic Resonance Imaging. CT: Computerized tomography. US: Ultrasound. mIBG: meta-iodo-benzyl-guanidine scintigraphy. BM: Medullar Bone exploration. Treatments: CBCA: Carboplatine, VP16: Etoposide, VCR: Vincristine, CDDP: Cisplatin. MAT: Myeloablative therapy. PBSC: Peripherical Blood Stem Cell. Bu Mel: Busulphan Mephalan. Rx: Radiotherapy.

**Figure S2:**
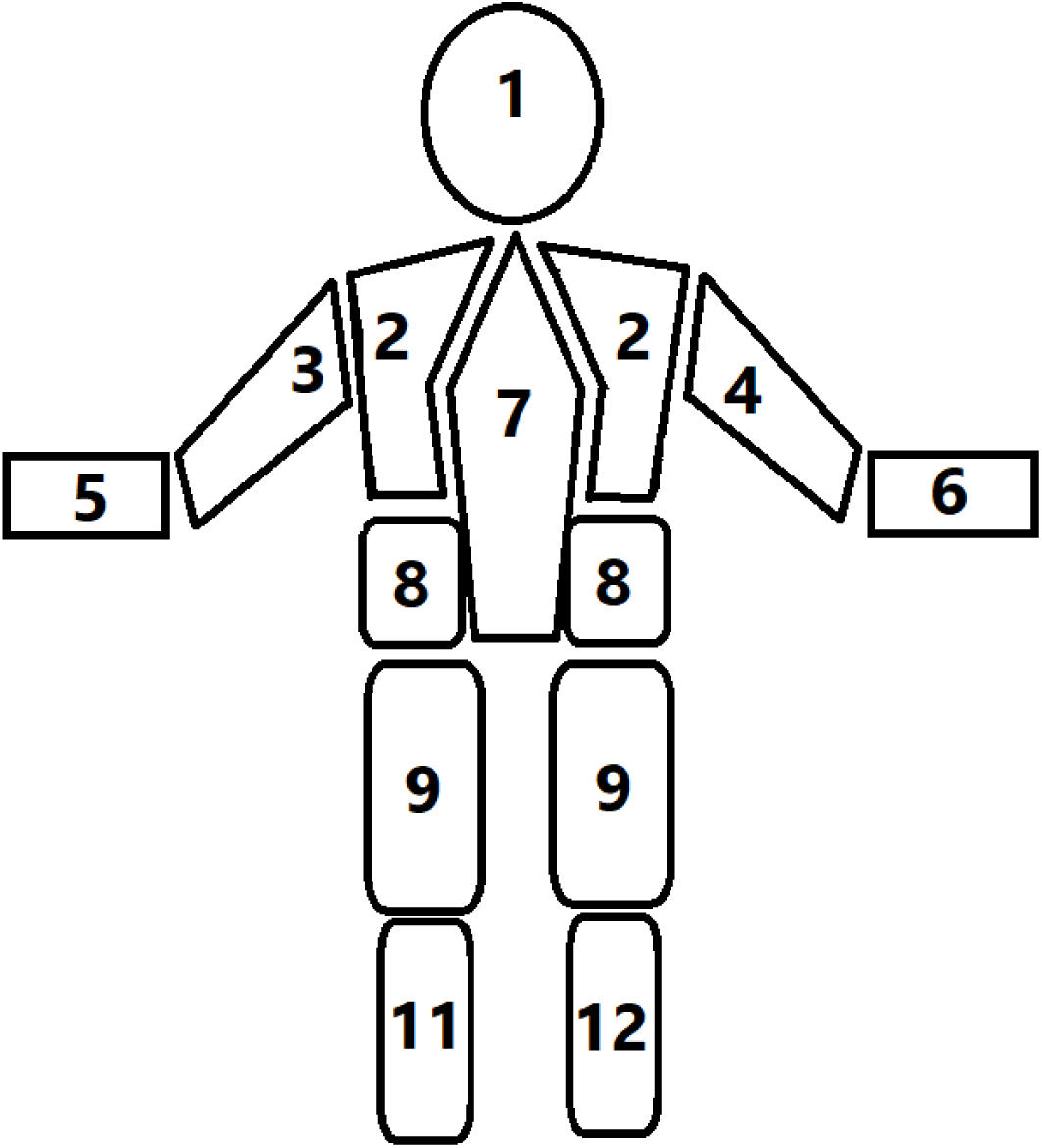
SIOPEN scoring. To score patients, the skeleton is divided into 12 segments, and for each of them extension of the lesions is scored as: - 0: no lesion
- 1 for 1 lesion
- 2 for 2 lesions
- 3 for 3 lesions
- 4 for > 3 lesions but < 50% of the concerned segment
- 5 for diffuse disease but < 95% of the whole segment
- 6 for difsuse disease > 95% of whole segment The SIOPEN score is then defined as the sum of each segment’s score.

**Figure S3:**
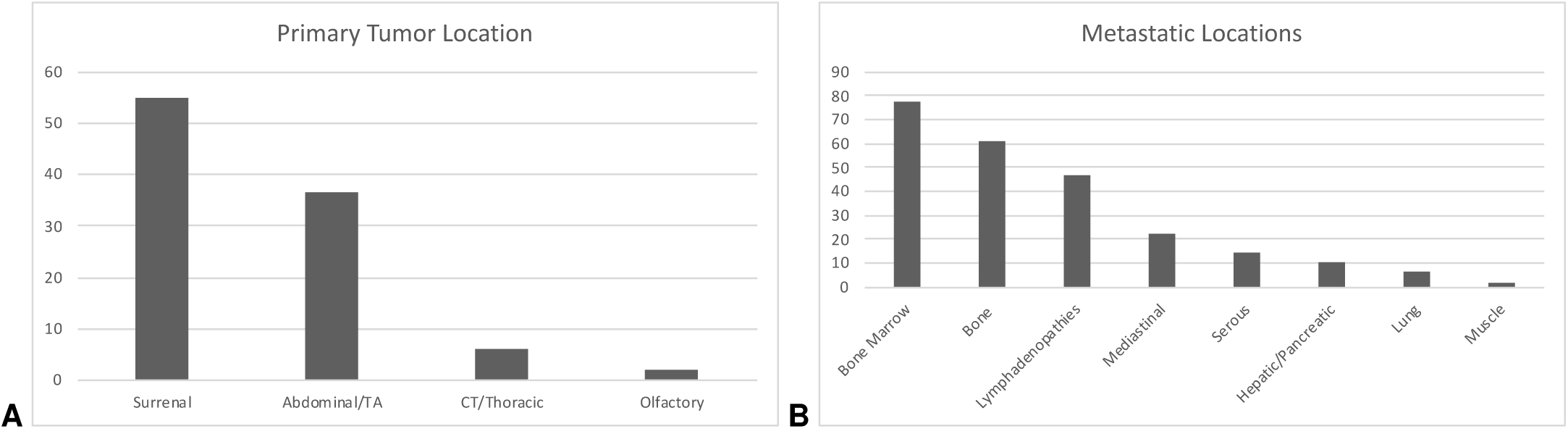
Primary tumor and metastases location.

**Figure S4:**
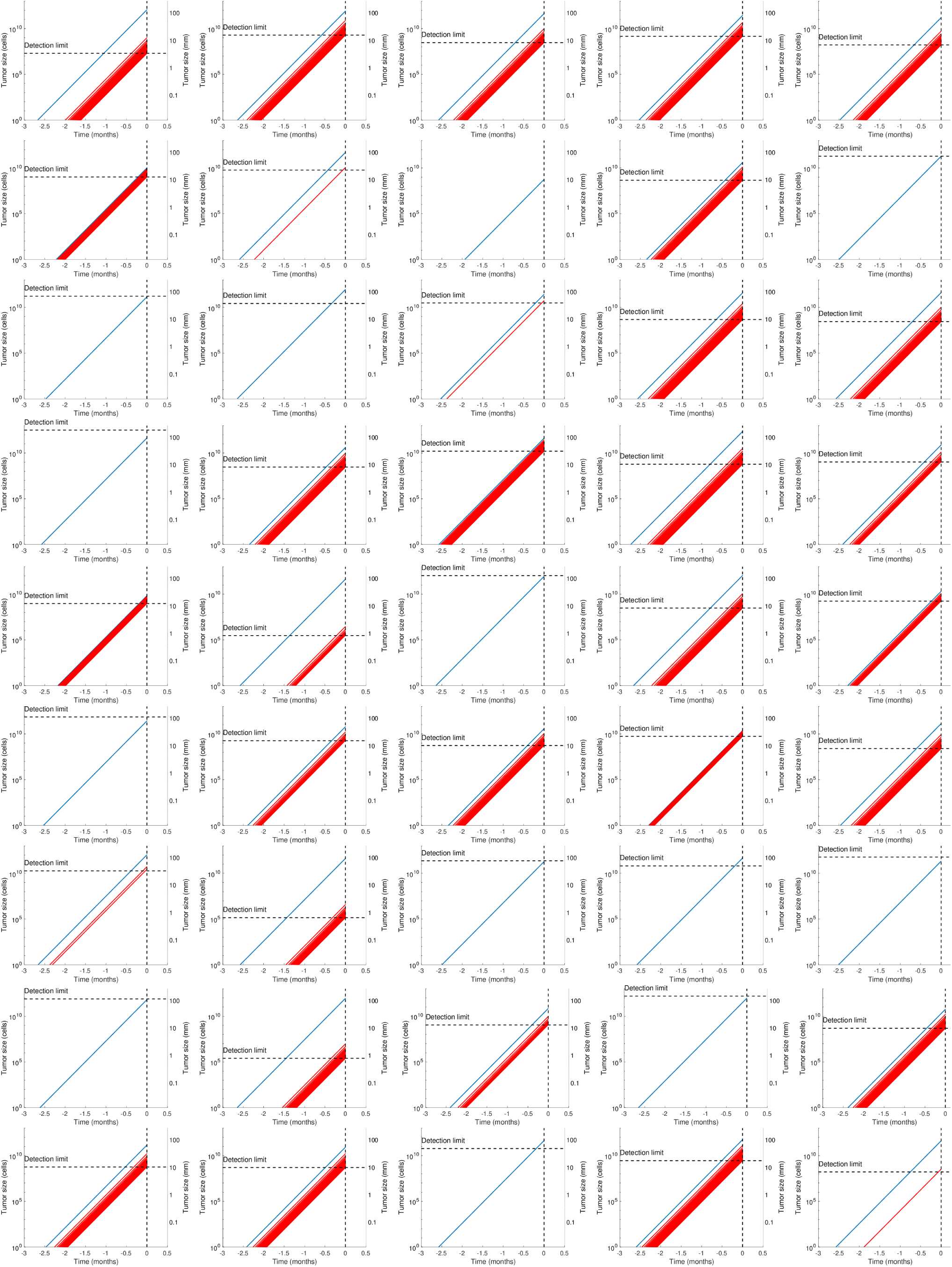
Simulations of all patients.

**Figure S5:**
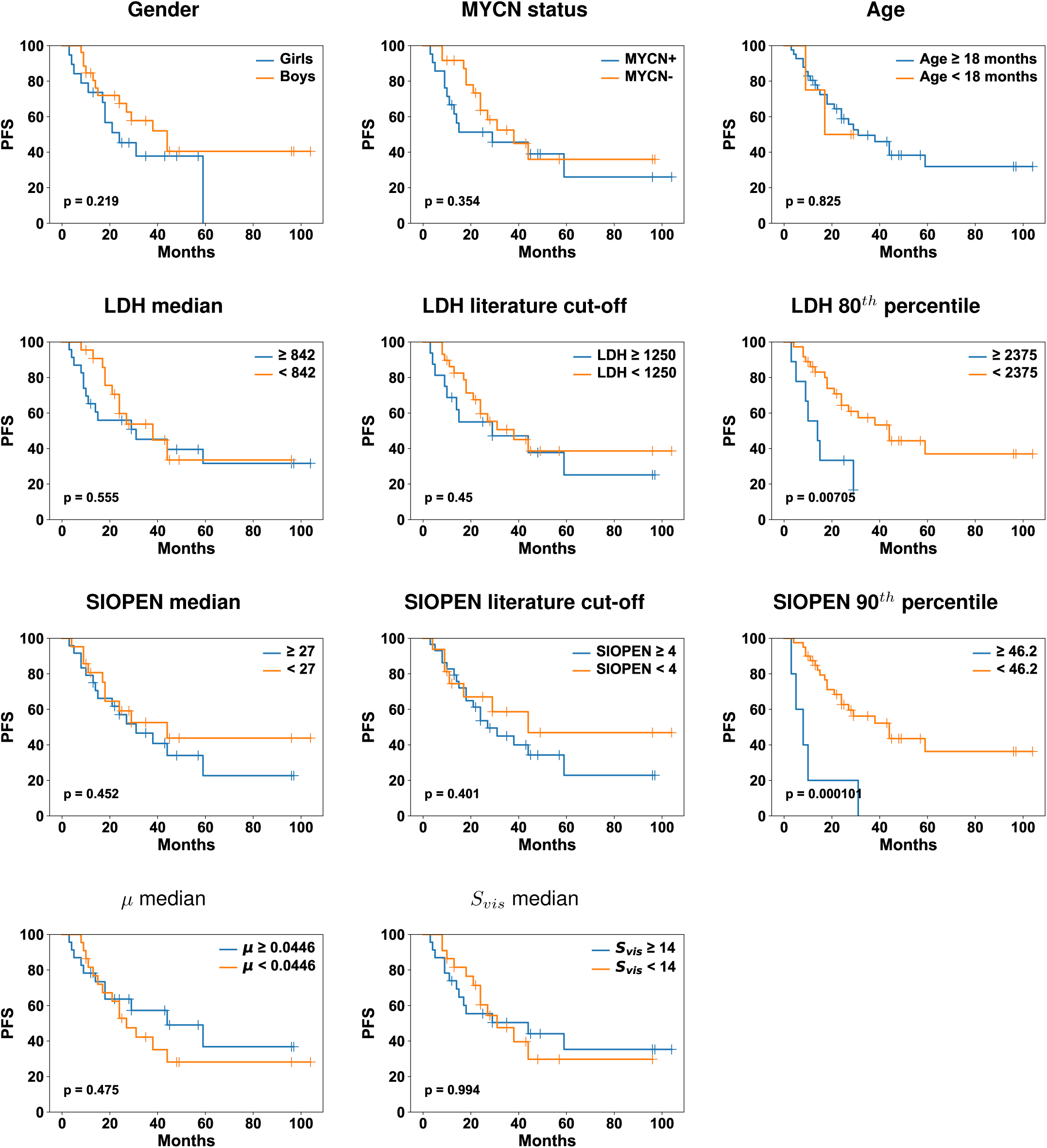
PFS analysis in dichotomized groups.

**Figure S6:**
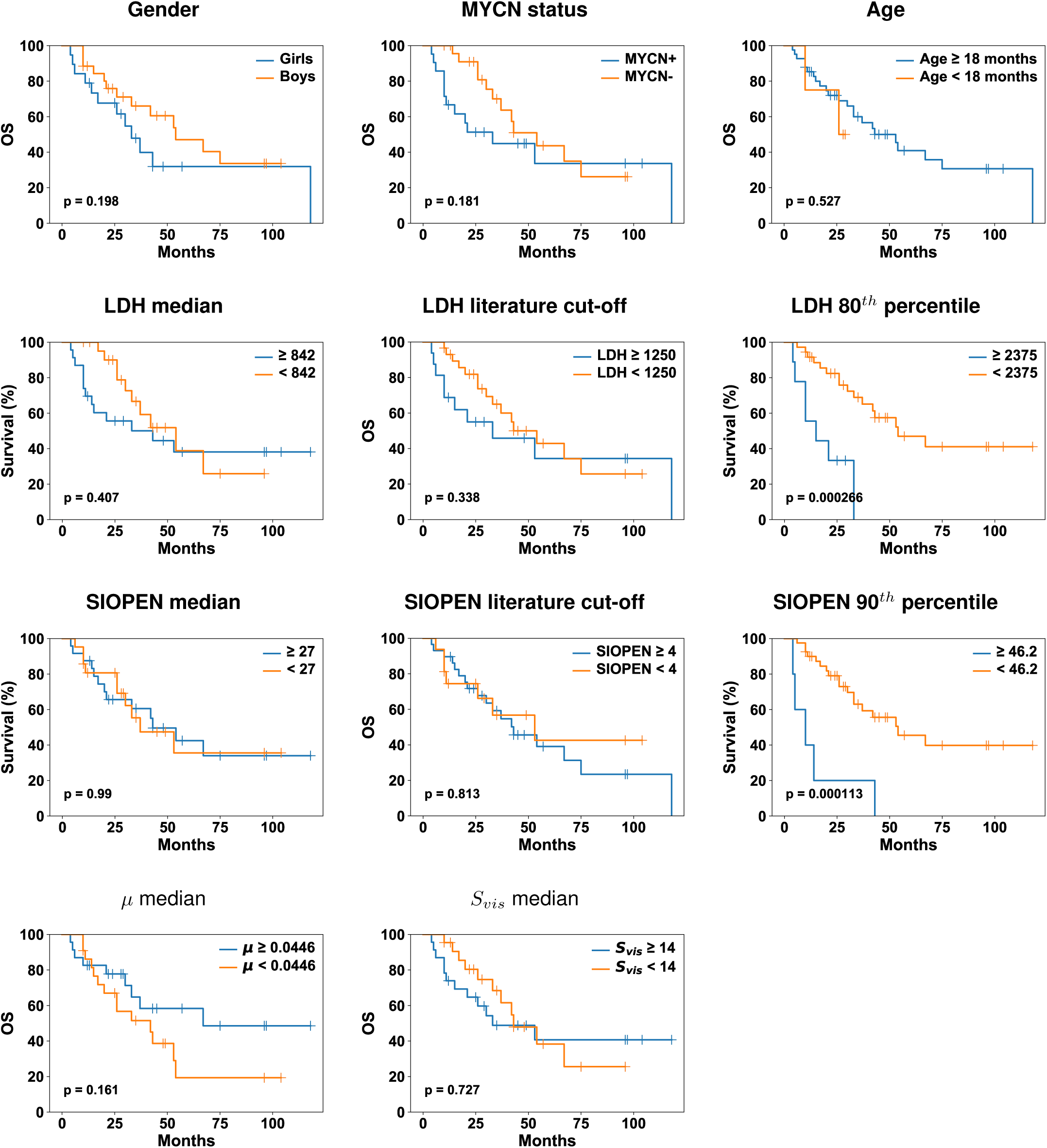
OS analysis in dichotomized groups.

**Figure S7:**
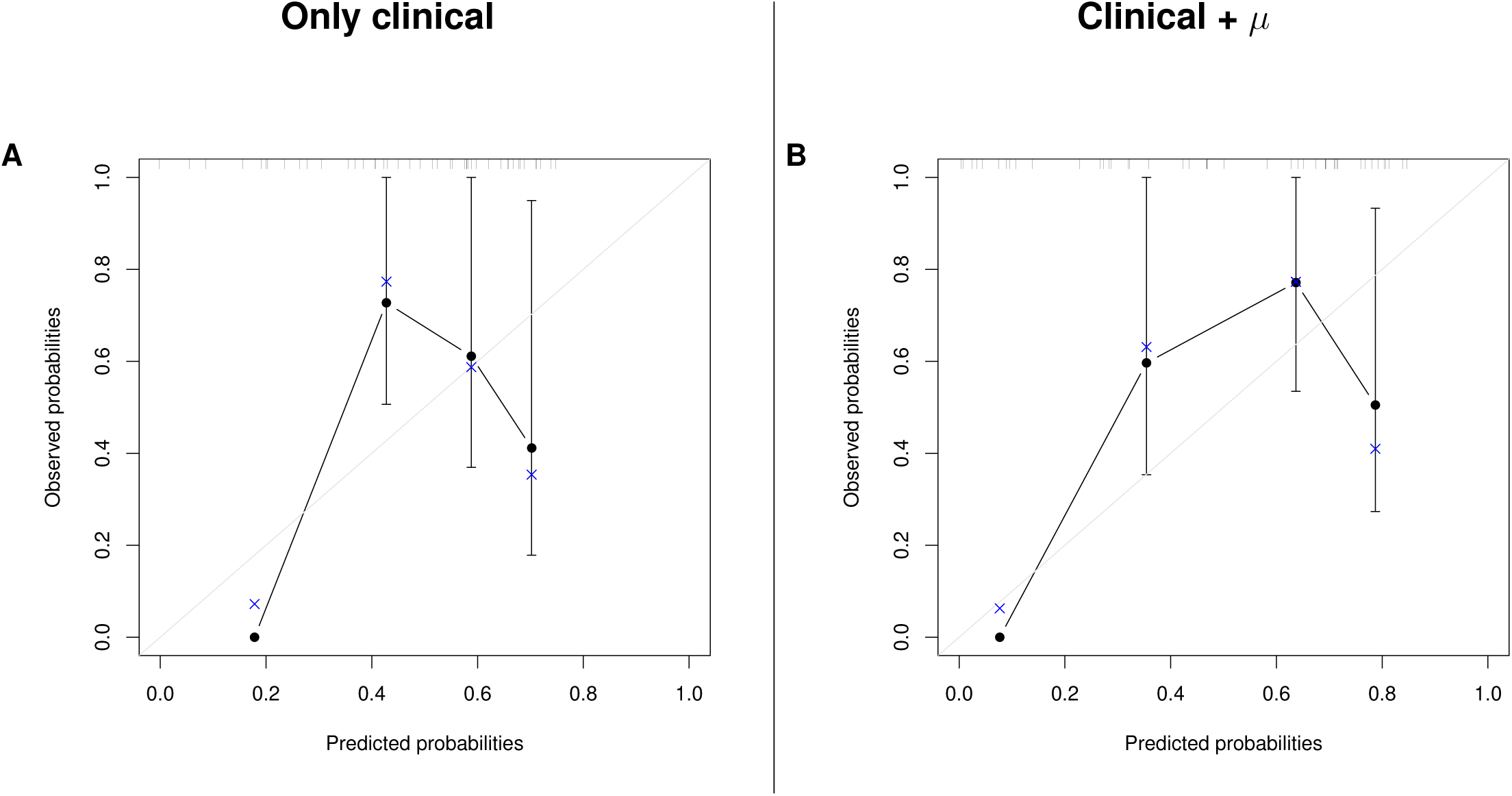
Calibration plots.

## Notes

**Conflict of interest:** The authors declare no potential conflicts of interest

### Competing Interest Statement

The authors have declared no competing interest.

### Funding Statement

No external funding was received

## References

1. Siegel RL, Miller KD, Jemal A. Cancer statistics, 2020. CA: A Cancer Journal for Clinicians. 2020;70:7–30.

2. Ahmed AA, Zhang L, Reddivalla N, Hetherington M. Neuroblastoma in children: Update on clinicopathologic and genetic prognostic factors. Pediatr Hematol Oncol. 2017;34:165–85.

3. Cohn SL, Pearson ADJ, London WB, Monclair T, Ambros PF, Brodeur GM, et al. The International Neuroblastoma Risk Group (INRG) classification system: an INRG Task Force report. J Clin Oncol. 2009;27:289–97.

4. Cheung N-KV, Dyer MA. Neuroblastoma: Developmental Biology, Cancer Genomics, and Immunotherapy. Nat Rev Cancer. 2013;13:397–411.

5. Whittle SB, Smith V, Doherty E, Zhao S, McCarty S, Zage PE. Overview and recent advances in the treatment of neuroblastoma. Expert Review of Anticancer Therapy. 2017;17:369–86.

6. Sokol E, Desai AV. The Evolution of Risk Classification for Neuroblastoma. Children (Basel). 2019;6.

7. London WB, Castleberry RP, Matthay KK, Look AT, Seeger RC, Shimada H, et al. Evidence for an age cutoff greater than 365 days for neuroblastoma risk group stratification in the Children’s Oncology Group. J Clin Oncol. 2005;23:6459–65.

8. Valteau-Couanet D, Schleiermacher G, Sarnacki S, Pasqualini C. [High-risk neuroblastoma treatment strategy: The experience of the SIOPEN group]. Bull Cancer. 2018;105:918–24.

9. Morgenstern DA, Pötschger U, Moreno L, Papadakis V, Owens C, Ash S, et al. Risk stratification of high-risk metastatic neuroblastoma: A report from the HR-NBL- 1/SIOPEN study. Pediatr Blood Cancer. 2018;65:e27363.

10. Matthay KK, Reynolds CP, Seeger RC, Shimada H, Adkins ES, Haas-Kogan D, et al. Long-term results for children with high-risk neuroblastoma treated on a randomized trial of myeloablative therapy followed by 13-cis-retinoic acid: a children’s oncology group study. J Clin Oncol. 2009;27:1007–13.

11. Ladenstein R, Pötschger U, Pearson ADJ, Brock P, Luksch R, Castel V, et al. Busulfan and melphalan versus carboplatin, etoposide, and melphalan as high-dose chemotherapy for high-risk neuroblastoma (HR-NBL1/SIOPEN): an international, randomised, multi-arm, open-label, phase 3 trial. Lancet Oncol. 2017;18:500–14.

12. London WB, Bagatell R, Weigel BJ, Fox E, Guo D, Van Ryn C, et al. Historical time to disease progression and progression-free survival in patients with recurrent/refractory neuroblastoma treated in the modern era on Children’s Oncology Group early-phase trials. Cancer. 2017;123:4914–23.

13. Basta NO, Halliday GC, Makin G, Birch J, Feltbower R, Bown N, et al. Factors associated with recurrence and survival length following relapse in patients with neuroblastoma. Br J Cancer. 2016;115:1048–57.

14. Skipper HE, Schabel FM, Wilcox WS. Experimental evaluation of potential anticancer agents XIII. On the criteria and kinetics associated with “curability” of experimental leukemia. Cancer Chemother Rep. 1964;35:1–111.

15. Altrock PM, Liu LL, Michor F. The mathematics of cancer: integrating quantitative models. Nat Rev Cancer. 2015;15:730–45.

16. Benzekry S, Pasquier E, Barbolosi D, Lacarelle B, Barlési F, André N, et al. Metronomic reloaded: Theoretical models bringing chemotherapy into the era of precision medicine. Semin Cancer Biol. 2015;35:53–61.

17. Barbolosi D, Ciccolini J, Lacarelle B, Barlési F, André N. Computational oncology--mathematical modelling of drug regimens for precision medicine. Nat Rev Clin Oncol. 2016;13:242–54.

18. Clancy CE, An G, Cannon WR, Liu Y, May EE, Ortoleva P, et al. Multiscale Modeling in the Clinic: Drug Design and Development. Ann Biomed Eng. 2016;44:2591–610.

19. Rajkomar A, Dean J, Kohane I. Machine Learning in Medicine. N Engl J Med. 2019;380:1347–58.

20. Topol EJ. High-performance medicine: the convergence of human and artificial intelligence. Nat Med. 2019;25:44.

21. van ‘t Veer LJ, Dai H, van de Vijver MJ, He YD, Hart AAM, Mao M, et al. Gene expression profiling predicts clinical outcome of breast cancer. Nature. 2002;415:530–6.

22. McKinney SM, Sieniek M, Godbole V, Godwin J, Antropova N, Ashrafian H, et al. International evaluation of an AI system for breast cancer screening. Nature. 2020;577:89–94.

23. Bruno R, Mercier F, Claret L. Evaluation of tumor size response metrics to predict survival in oncology clinical trials. Clin Pharmacol Ther. 2014;95:386–93.

24. Baratchart E, Benzekry S, Bikfalvi A, Colin T, Cooley LS, Pineau R, et al. Computational Modelling of Metastasis Development in Renal Cell Carcinoma. PLoS Comput Biol. 2015;11:e1004626.

25. Bilous M, Serdjebi C, Boyer A, Tomasini P, Pouypoudat C, Barbolosi D, et al. Quantitative mathematical modeling of clinical brain metastasis dynamics in non-small cell lung cancer. Sci Rep. 2019;9:13018.

26. Benzekry S, Tracz A, Mastri M, Corbelli R, Barbolosi D, Ebos JML. Modeling Spontaneous Metastasis following Surgery: An In Vivo-In Silico Approach. Cancer Res. 2016;76:535–47.

27. Nicolo C, Perier C, Prague M, MacGrogan G, Saut O, Benzekry S. Machine learning versus mechanistic modeling for prediction of metastatic relapse in breast cancer. JCO Clin Cancer Inform. 2020;in press.

28. Pang QM, Li K, Ma LJ, Sun RP. Clinical research on neuroblastoma based on serum lactate dehydrogenase. J Biol Regul Homeost Agents. 2015;29:131–4.

29. Dorneburg C, Fischer M, Barth TFE, Mueller-Klieser W, Hero B, Gecht J, et al. LDHA in Neuroblastoma Is Associated with Poor Outcome and Its Depletion Decreases Neuroblastoma Growth Independent of Aerobic Glycolysis. Clin Cancer Res. 2018;24:5772–83.

30. Shulkin BL, Shapiro B. Current concepts on the diagnostic use of MIBG in children. J Nucl Med. 1998;39:679–88.

31. Matthay KK, Shulkin B, Ladenstein R, Michon J, Giammarile F, Lewington V, et al. Criteria for evaluation of disease extent by 123I-metaiodobenzylguanidine scans in neuroblastoma: a report for the International Neuroblastoma Risk Group (INRG) Task Force. Br J Cancer. 2010;102:1319–26.

32. Ara T, DeClerck YA. Mechanisms of invasion and metastasis in human neuroblastoma. Cancer Metastasis Rev. 2006;25:645–57.

33. Bleeker G, Tytgat GAM, Adam JA, Caron HN, Kremer LCM, Hooft L, et al. 123I-MIBG scintigraphy and 18F-FDG-PET imaging for diagnosing neuroblastoma. Cochrane Database Syst Rev. 2015;CD009263.

34. Ladenstein R, Lambert B, Pötschger U, Castellani M-R, Lewington V, Bar-Sever Z, et al. Validation of the mIBG skeletal SIOPEN scoring method in two independent high-risk neuroblastoma populations: the SIOPEN/HR-NBL1 and COG-A3973 trials. Eur J Nucl Med Mol Imaging. 2018;45:292–305.

35. Park JR, Bagatell R, Cohn SL, Pearson AD, Villablanca JG, Berthold F, et al. Revisions to the International Neuroblastoma Response Criteria: A Consensus Statement From the National Cancer Institute Clinical Trials Planning Meeting. J Clin Oncol. 2017;35:2580–7.

36. Iwata K, Kawasaki K, Shigesada N. A dynamical model for the growth and size distribution of multiple metastatic tumors. J Theor Biol. 2000;203:177–86.

37. Hartung N. Efficient resolution of metastatic tumor growth models by reformulation into integral equations. Discrete Contin Dyn Syst Ser B. 2015;20:445–67.

38. Spratt JS, Meyer JS, Spratt JA. Rates of growth of human solid neoplasms: Part I. J Surg Oncol. 1995;60:137–46.

39. Harrell FE, Lee KL, Mark DB. Multivariable prognostic models: issues in developing models, evaluating assumptions and adequacy, and measuring and reducing errors. Stat Med. 1996;15:361–87.

40. Gallik KL, Treffy RW, Nacke LM, Ahsan K, Rocha M, Green-Saxena A, et al. Neural crest and cancer: Divergent travelers on similar paths. Mech Dev. 2017;148:89–99.

41. Borriello L, Seeger RC, Asgharzadeh S, DeClerck YA. More than the genes, the tumor microenvironment in neuroblastoma. Cancer Lett. 2016;380:304–14.

42. Wilkie KP, Hahnfeldt P. Tumor-immune dynamics regulated in the microenvironment inform the transient nature of immune-induced tumor dormancy. Cancer Res. 2013;73:3534–44.

43. Vanichapol T, Chutipongtanate S, Anurathapan U, Hongeng S. Immune Escape Mechanisms and Future Prospects for Immunotherapy in Neuroblastoma. Biomed Res Int. 2018;2018:1812535.

44. Jubierre L, Jiménez C, Rovira E, Soriano A, Sábado C, Gros L, et al. Targeting of epigenetic regulators in neuroblastoma. Exp Mol Med. 2018;50:51.

45. Benzekry S, André N, Benabdallah A, Ciccolini J, Faivre C, Hubert F, et al. Modelling the impact of anticancer agents on metastatic spreading. Math Model Nat Phenom. 2012;7:306–36.

46. Benzekry S, Lamont C, Barbolosi D, Hlatky L, Hahnfeldt P. Mathematical Modeling of Tumor-Tumor Distant Interactions Supports a Systemic Control of Tumor Growth. Cancer Res. 2017;77:5183–93.

47. Ciccolini J, Barbolosi D, Meille C, Lombard A, Serdjebi C, Giacometti S, et al. Pharmacokinetics and Pharmacodynamics-Based Mathematical Modeling Identifies an Optimal Protocol for Metronomic Chemotherapy. Cancer Res. 2017;77:4723–33.

48. He Y, Kodali A, Wallace DI. Predictive Modeling of Neuroblastoma Growth Dynamics in Xenograft Model After Bevacizumab Anti-VEGF Therapy. Bull Math Biol. 2018;80:2026–48.

49. Kasemeier-Kulesa JC, Schnell S, Woolley T, Spengler JA, Morrison JA, McKinney MC, et al. Predicting neuroblastoma using developmental signals and a logic-based model. Biophys Chem. 2018;238:30–8.

50. Hidalgo MR, Amadoz A, Çubuk C, Carbonell-Caballero J, Dopazo J. Models of cell signaling uncover molecular mechanisms of high-risk neuroblastoma and predict disease outcome. Biol Direct. 2018;13:16.

51. Zhang L, Lv C, Jin Y, Cheng G, Fu Y, Yuan D, et al. Deep Learning-Based Multi-Omics Data Integration Reveals Two Prognostic Subtypes in High-Risk Neuroblastoma. Front Genet. 2018;9:477.

52. Depuydt P, Koster J, Boeva V, Hocking TD, Speleman F, Schleiermacher G, et al. Meta-mining of copy number profiles of high-risk neuroblastoma tumors. Sci Data. 2018;5:180240.

53. Salazar BM, Balczewski EA, Ung CY, Zhu S. Neuroblastoma, a Paradigm for Big Data Science in Pediatric Oncology. Int J Mol Sci. 2016;18.

54. Vaghi C, Rodallec A, Fanciullino R, Ciccolini J, Mochel JP, Mastri M, et al. Population modeling of tumor growth curves and the reduced Gompertz model improve prediction of the age of experimental tumors. PLOS Computational Biology. Public Library of Science; 2020;16:e1007178.

